# Impacts of climate change on diarrhoeal disease hospitalisations: how does the Global Warming Targets of 1.5 - 2°C affect Dhaka, Bangladesh?

**DOI:** 10.1101/2024.04.10.24305607

**Authors:** Farhana Haque, Fiona C Lampe, Shakoor Hajat, Katerina Stavrianaki, S. M. Tafsir Hasan, ASG Faruque, Tahmeed Ahmed, Shamim Jubayer, Ilan Kelman

## Abstract

Dhaka is one of the world’s densely populated cities and faces significant public health challenges including high burden of diarrhoeal diseases. Climate change is intensifying existing environmental problems including urban heat island effect and poor water quality. While numerous epidemiological studies have linked meteorological factors to diarrhoeal diseases in Bangladesh, assessment of the impacts of future climate change on diarrhoeal diseases is scarce. We provide the assessment of climate change impacts on diarrhoeal disease in Dhaka and project future health risks under climate change scenarios. About 3 million acute diarrhoea cases presenting to the Dhaka Hospital of the International Centre for Diarrhoeal Disease Research Bangladesh (icddr,b) during 1981 - 2010 were linked to daily temperature, rainfall and humidity and association investigated using time series adapted negative binomial regression models employing constrained distributed lag linear models. The findings were applied to climate projections to estimate future risks of diarrhoea under various global warming scenarios. There was a significantly raised risk of diarrhoea hospitalisation in all ages with daily mean temperature (RR: 3.4, 95% CI: 3.0 – 3.7) after controlling for the confounding effects of heavy rainfall, humidity, autocorrelations, day of the week effect, long-term time, and seasonal trends. Using the incidence rate ratio (IRR) of 1.034, temperature increases based on the global warming targets of 1.5 – 2°C could result in an increase of diarrhoea hospitalisations by 4.5 – 7.4% in all age groups by the 2100s. These effects were more pronounced among <5 children where the predicted temperature increases could raise diarrhoea hospitalisation by 5.7% - 9.4%. Diarrhoea hospitalisation will increase significantly in Dhaka even if the global warming targets adopted by the Paris Agreement is reached. This underscores the importance of preparing the city for management and prevention of diarrhoeal diseases.

## Introduction

The global mean surface temperature is projected to rise by 2.6°C–4.8°C under the high emissions scenarios in 2100 even if all countries reduce their greenhouse gas emissions as pledged in the Paris Agreement [1]. Such temperature increase poses significant threat to the human society as global warming can adversely affect human health and wellbeing, particularly in the low- and middle-income countries (LMICs) that have little economic capacity to take adequate mitigation and adaptation measures [2–4]. Climate change is projected to exert significant impact on hydrological systems, water availability and water quality. As a result, waterborne diseases, particularly diarrhoeal diseases are expected to be adversely affected by the impacts of climate change [5, 6]. Diarrhoeal diseases, with an estimated annual global incidence of 2.39 billion episodes and 1.3 million deaths in all ages according to the global burden of disease study in 2015, remain one of the leading causes of morbidity and mortality worldwide [7]. Given the high baseline rates, even a small increase in risk stemming from climate change can significantly increase public health burdens in the LMICs.

Despite the concerns that climate change can cause excess diarrhoea, only a limited number of studies have attempted to quantify the effects of climate change on diarrhoea across the globe with variable results [8–10]. Using projections from one climate model and empirical data from two studies conducted in Fiji [11] and Peru [12], one study inferred that global warming of 1°C by the 2030s will lead to an increase in diarrhoea by 0–10% [8]. Another study projected increases of 8–11% in relative risks of diarrhoea by 2010 – 2039 using data from five empirical studies across the globe and 19-member climate model ensemble [9]. On the other hand, Moors et al. (2013) calculated increases of 0–21% in diarrhoea by the 2040s, using the latest regional climate projections for northern India [10]. A recent study projected over a 10% rise in heat-related diarrhoeal diseases in the Gaza Strip, Palestine under the 2 °C global warming scenario [13].

Bangladesh is a low- and middle-income country (LMIC) in Southeast Asia with a low-lying topography, poor infrastructure and high population density [14]. Bangladesh has been classified in the Sixth Assessment Report of the Intergovernmental Panel on Climate Change (IPCC) as a country that is highly vulnerable to the adverse impacts of climate change [1]. As temperatures continue to rise and rainfall becomes more erratic, water stress and food insecurity may increase in Bangladesh [14]. Though Bangladesh has made strong progress towards ensuring better quality drinking water—the switching to improved water infrastructure—much of what is tapped remains contaminated with dangerous microbes, heavy metals, or salt. Moreover, infrastructure alone may not ensure quality of drinking water supplies at all times [15]. Diarrhoeal disease is endemic in Bangladesh. While the exact prevalence estimate of diarrhoea is lacking in Bangladesh, diarrhoea is a major cause of morbidity in the country [7]. According to the Bangladesh Demographic and Health Surveys (BDHS), the reported prevalence of diarrhoea in the previous two weeks among under-five children was 5.7% and the prevalence of diarrhoea with blood was 1% in 2014. Despite meeting the water MDG target successfully in 2010, outbreaks and hyper-endemicity of diarrhoeal diseases continue to plague the developing nations including Bangladesh [16]. With more than an estimated 76 million people affected with diarrhoeal disease episodes in all age groups in Bangladesh during 2015 [7], the potential impact of long term variability in the climate on the incidence of diarrhoeal disease in the future could be concerning for Bangladesh.

This study aims to assess the impacts of meteorological factors on acute diarrhoeal disease in Dhaka, Bangladesh using daily time-series datasets and quantify the effects of temperature increase under future climate change scenarios.

## Materials and methods

### Study setting

Dhaka, located at 23°42′N 90°22′E on the eastern banks of the Buriganga River, is the capital and largest city of Bangladesh. Tropical vegetation and moist soils characterize the land, which is flat and close to sea level. Dhaka, a low-lying, predominantly coastal city surrounded by five major rivers and 50 water channels, is at risk of flooding from rising sea levels [17]. Several dykes and embankments and interim boundaries created to protect Dhaka from flooding together with inadequate drainage and water pumping systems frequently results in water logging of the protected city area during the monsoons [18]. With a density of 23,234 people per square kilometres within a total area of 300 square kilometres of the city area, Dhaka city faces a number of environmental and societal challenges including urban heat island effect, insufficient infrastructure, poor water quality, inadequate sanitation, inadequate waste management, poor hygiene brought about by poverty [19, 20].

### Health data

Data on the daily number of acute diarrhoea cases (defined as patients passing ≥3 loose stools per 24h due to any cause) presenting to the Dhaka Hospital of the International Centre for Diarrhoeal Diseases Research, Bangladesh (icddr,b) between 1981–2010 were obtained on 7 October 2020. The Dhaka hospital is the largest diarrhoeal disease hospital that predominantly served an urban population of approximately 14.7 million and provided free treatment to more than 140,000 patients with diarrhoea in 2010 [21]. Given that reliable records of the total number of patients with acute diarrhoea admitted per day or their disease onset dates were not available for the study period (1981–2010), information from the Diarrhoeal Disease Surveillance System (DDSS) was obtained instead to estimate the total number of patients hospitalised with diarrhoea per day. The DDSS was established in 1979 to record the information of a sample of acute diarrhoea patients who sought care from the icddr,b Dhaka Hospital [22]. During 1981–1995, the DDSS enrolled every 25th patient and during 1996–2010, the DDSS enrolled every 50th patient who were admitted in the icddr,b Dhaka Hospital into the surveillance system. The Dhaka Hospital began keeping electronic records of all patients seeking care since 2010 [23]. We did not access any information that could identify individual participants during or after data collection. We did not collect any data beyond the study period. Additional information on health data is provided in the supplementary material (S1 File).

### Meteorological data

Daily climate parameters including mean, maximum and minimum temperature, cumulative rainfall, and relative humidity between 1 January 1980 and 31 December 2010 for Dhaka City were collected from the Bangladesh Meteorological Department (BMD). The BMD maintains 3-hourly records of temperature data from three validated weather stations across Dhaka. All data are publicly accessible at https://bmd.gov.bd/.

While climate projections for Dhaka city are limited, a study projected an increase in maximum temperature by 1.3 – 4.3°C in Bangladesh for different representative concentration pathway (RCP) scenarios using the Coupled Model Intercomparison Project Phase 5 (CMIP5) global climate models (GCMs) models by the 2100 [24]. Various regional climate models (RCMs) using different IPCC greenhouse gas emission scenarios predicted a likely increase in the mean temperature by 1 – 3°C during 2030 – 2060 in Bangladesh [25]. Another study projected change of mean surface air temperature in different months by 0.5 – 2.1°C and 0.9 – 3.5°C for the year 2050 and 2060, respectively [26]. For the SRES A1B emissions scenario (representing a future world of very rapid economic growth, low population growth and rapid introduction of new and more efficient technology), the projected temperature increases over the entire country are in the region of 3 to 3.5°C by the 2100s according to the UK’s Met Office Hadley Centre [27]. On the other hand, a study based on an ensemble of projections using the high resolution (10 km) MIT Regional Climate Model (MRCM) projected an increase of 1.2 – 1.3°C in daily maximum temperature by the 2050s for Bangladesh compared to a reference period of 1976 – 2005. Using statistical downscaling model, one study predicted an increase of 2.7°C and 2.1°C in average annual maximum and minimum temperature per century [28]. Using multi-model ensemble (MME) of 40 CMIP5 GCMs, a recent study showed that mean annual maximum temperature over Bangladesh will increase by 0.61-1.75°C in the near future (2021-2060) and 0.91 – 3.85°C in the far future (2071-2100) [14]. Given that different climate projections suggest a variable increase in daily temperature across Bangladesh ranging between 0.91 – 4.3°C by the 2100s and because climate projections are prone to biases [9], we estimated the increased risk of diarrhoea under various projected temperatures. In addition, we estimated the increase in relative risk of diarrhoea hospitalisation for the global warming targets agreed in the Paris Agreement since many countries have reaffirmed their commitment to hold the increase in global average temperature to 1.5 – 2°C above the pre-industrial levels.

### Additional data

Additional socio-economic data for sensitivity analyses were collected as follows: yearly total urban population projections for Dhaka was collected from the UN population prospects; the annual gross domestic product (GDP) per capita in Bangladesh was obtained from the Nationmaster website; literacy rate was obtained from the Bangladesh Bureau of Statistics and the 5-yearly data on access to improved water supply and sanitation were obtained from the Demographic and Health Surveillance (DHS) and the Joint Monitoring Programme (JMP). To compare how diarrhoea hospitalisation rates compared with national trends, we additionally collected annual diarrhoea hospitalisation and death data from the website of the Department of Health Services of the Ministry of Health and Family Welfare, Bangladesh (https://old.dghs.gov.bd/index.php/en/publications/health-bulletin/dghs-health-bulletin).

### Analysis

#### Epidemiological analysis to model baseline impacts

Negative binomial time-series regression models were used to assess acute associations between daily diarrhoea hospitalisation and daily ambient temperature, relative humidity and heavy rainfall (defined as the rainfall ≥95^th^ percentile for the study period). A series of linear-spline functions of time were used to flexibly model underlying long-term trends and seasonal patterns in the health data unrelated to climate factors [23, 29]. Seasonally adjusted negative binomial model was used to allow for overdispersion and model residuals were lagged by one day to adjust for the significant residual autocorrelation associated with the day before.

Distributed lag linear models (DLLMs) were then employed to model the delayed effects of each exposure that allowed the relationship to be simultaneously modelled at different lags of exposure. A maximum lag of 28 days was used to capture delayed effects. These models suggested a linear risk associated with mean temperature at lag 0-1 days, humidity at lag 0-3 days and heavy rainfall at lag 0-8 days. The cumulative effects of temperature, humidity or rainfall across all significant lags were therefore estimated using DLLMs. Although each variable was investigated separately, the final quantification was based on a model with all meteorological factors simultaneously entered in a constrained DLLM, which took the following form:

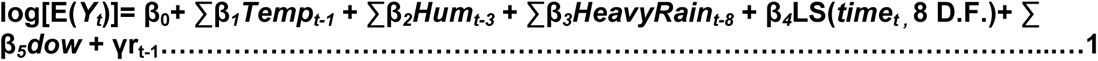

where *E(Y_t_)* is the expected diarrhoea hospitalisation count on day *t*; *Temp_t-1_* indicates the distributed lag of mean temperature at lags 0–1 days; and *Hum_t-3_* and *HeavyRain_t-8_* indicate the distributed lag of relative humidity and heavy rainfall at lags 0–3 and 0–8 days, respectively. *LS* indicates a linear spline of time with 8 degrees of freedom per year; *dow* is the categorical day of the week with a reference day of Friday; and *r_t-1_*is deviance residuals at 1 day lag obtained from the model without any autoregressive terms. The relative risk was estimated as incidence rate ratio (IRR) in the model and later converted into percentage.

Multiple sensitivity analyses were conducted by changing the amount of control for seasonality and long-term trends in the model (by using natural cubic splines and changing the number of knots in the spline-based approach). Further sensitivity analysis was conducted by including population of Dhaka into the model as an offset. The analysis was also repeated using the actual number of diarrheal disease patients enrolled into the DDSS during the study period. In addition, two separate models using a generalized estimating equations (GEE) approach were generated to investigate the potential modification by improved WASH interventions [30]. Diagnostic plots based on deviance residuals were generated for each model to check the model fit. The best model was chosen based on the lowest BIC value. All data were analysed using Stata/SE 18.0 (StataCorp, USA).

#### Estimating the impact of climate change on diarrhoea

The impact (I) of climate change on diarrhoea hospitalisation in the future was calculated by multiplying the projected increase in temperature (ΔT) with the percentage increase in relative risk (RR) of diarrhoea hospitalisations with each 1°C temperature increase using the formula below:

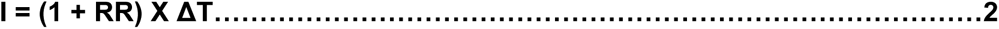

where RR is the increase in risk percentage of diarrhoea per 1°C increase in temperature and ΔT is the projected increase in temperature by the 2100s.

### Findings

During the 30-year study period between 1 January 1981 and 31 December 2010, approximately 2,983,850 cases of all ages presented to the icddr,b Dhaka Hospital for acute diarrhoeal disease. Among these, 58% were males and 54% were children aged <5 years. Figs 1 and 2 show the daily and monthly distributions of diarrhoea hospitalisation and mean temperature (°C) in Dhaka averaged across years 1981–2010. Diarrhoea hospitalisation for under 5 children peaked in April whereas diarrhoea hospitalisation for all ages revealed a large peak in April and a smaller peak in September (Fig 2).

**Fig 1.**
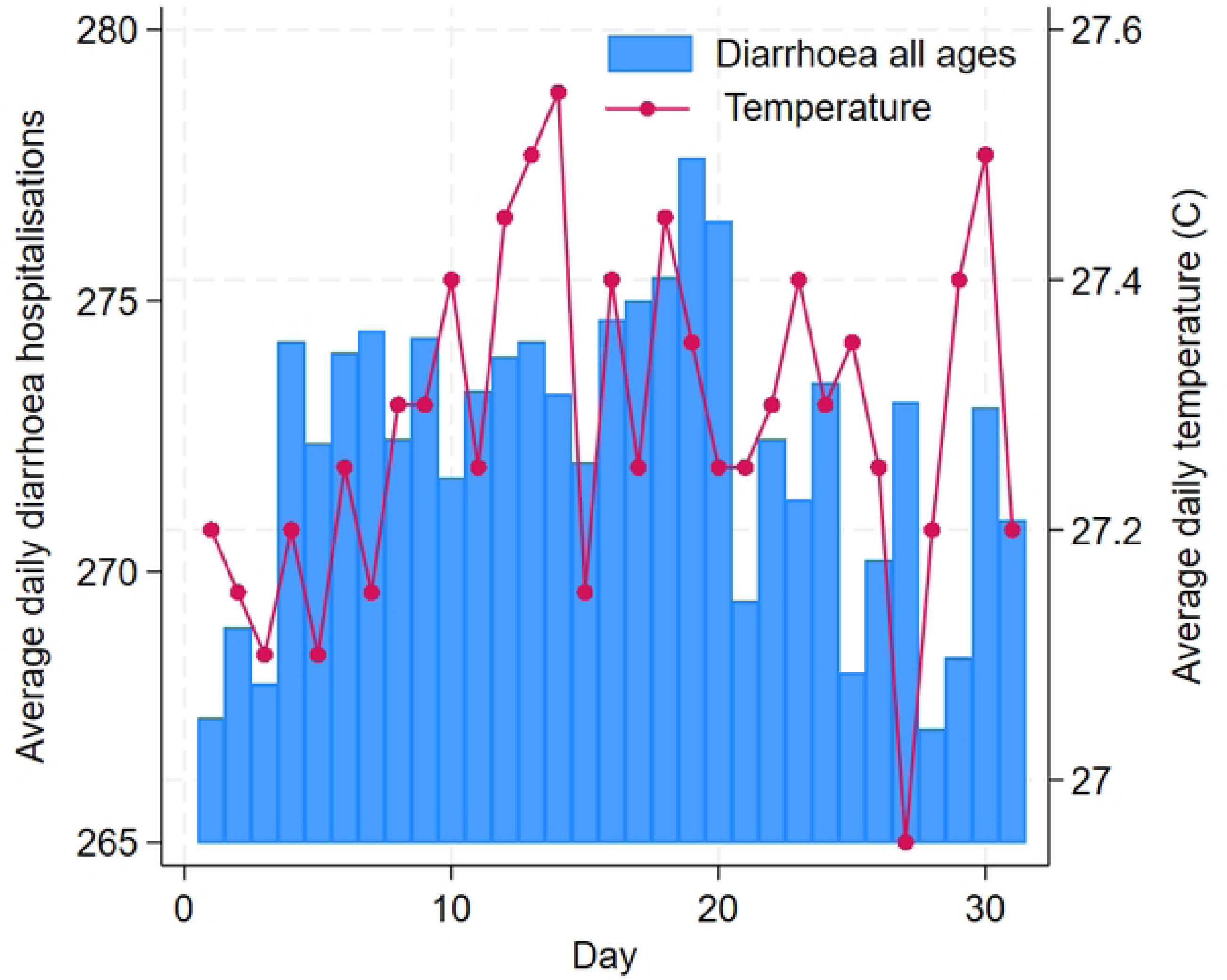
Daily distribution of diarrhoea hospitalisation and ambient temperature averaged across years 1981–2010.

**Fig 2.**
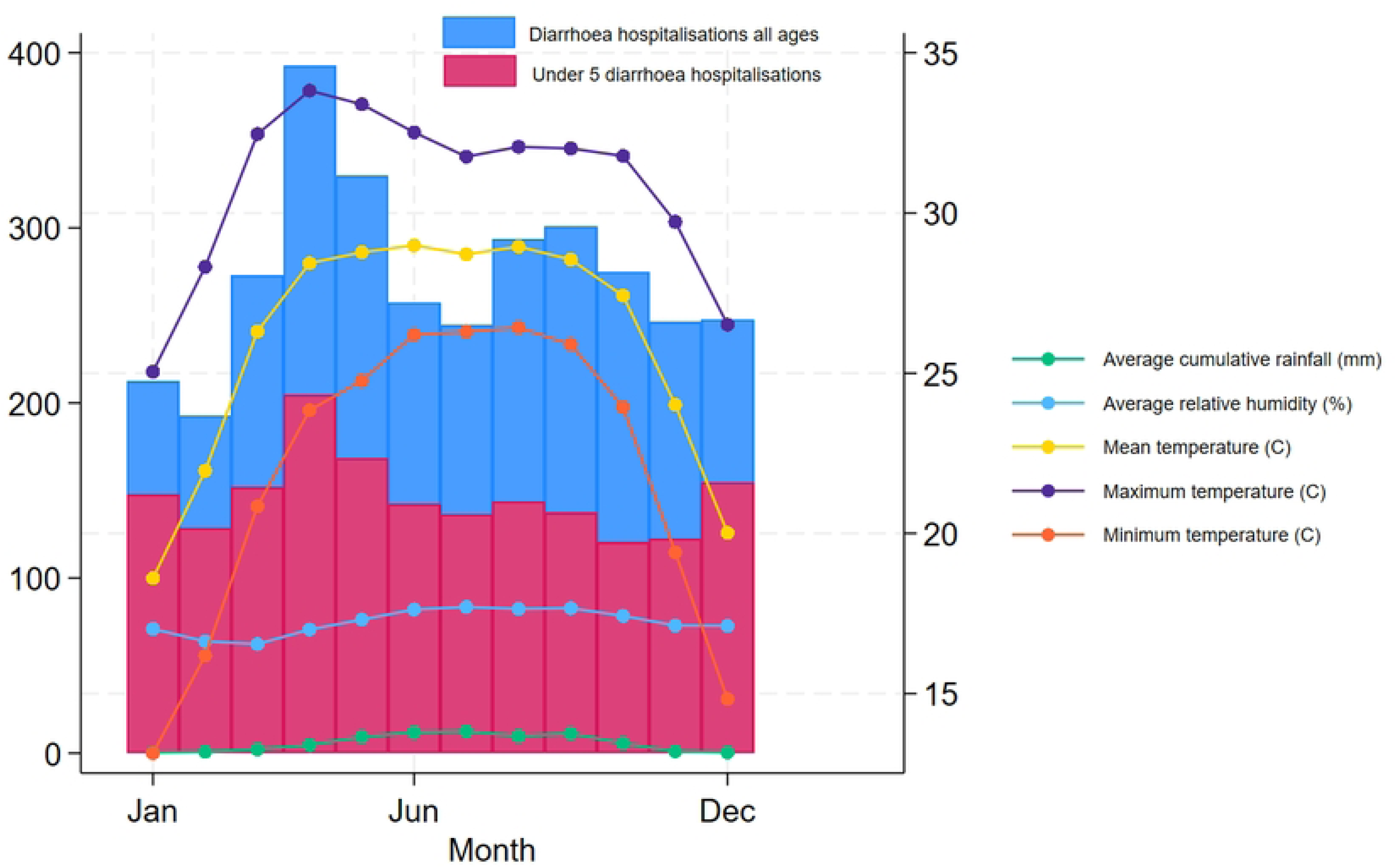
Monthly distribution of daily diarrhoea hospitalisation in all ages and ≤5 children and meteorological factors averaged across years 1981–2010.

According to Fig 3, hospitalisation rates in the Dhaka Hospital showed a declining trend with improvements in water supply, sanitation and socio-economic conditions. As evident from Figs 2 and 4, diarrhoea morbidity rates remain significantly high both in Dhaka and across Bangladesh during the study period.

**Fig 3.**
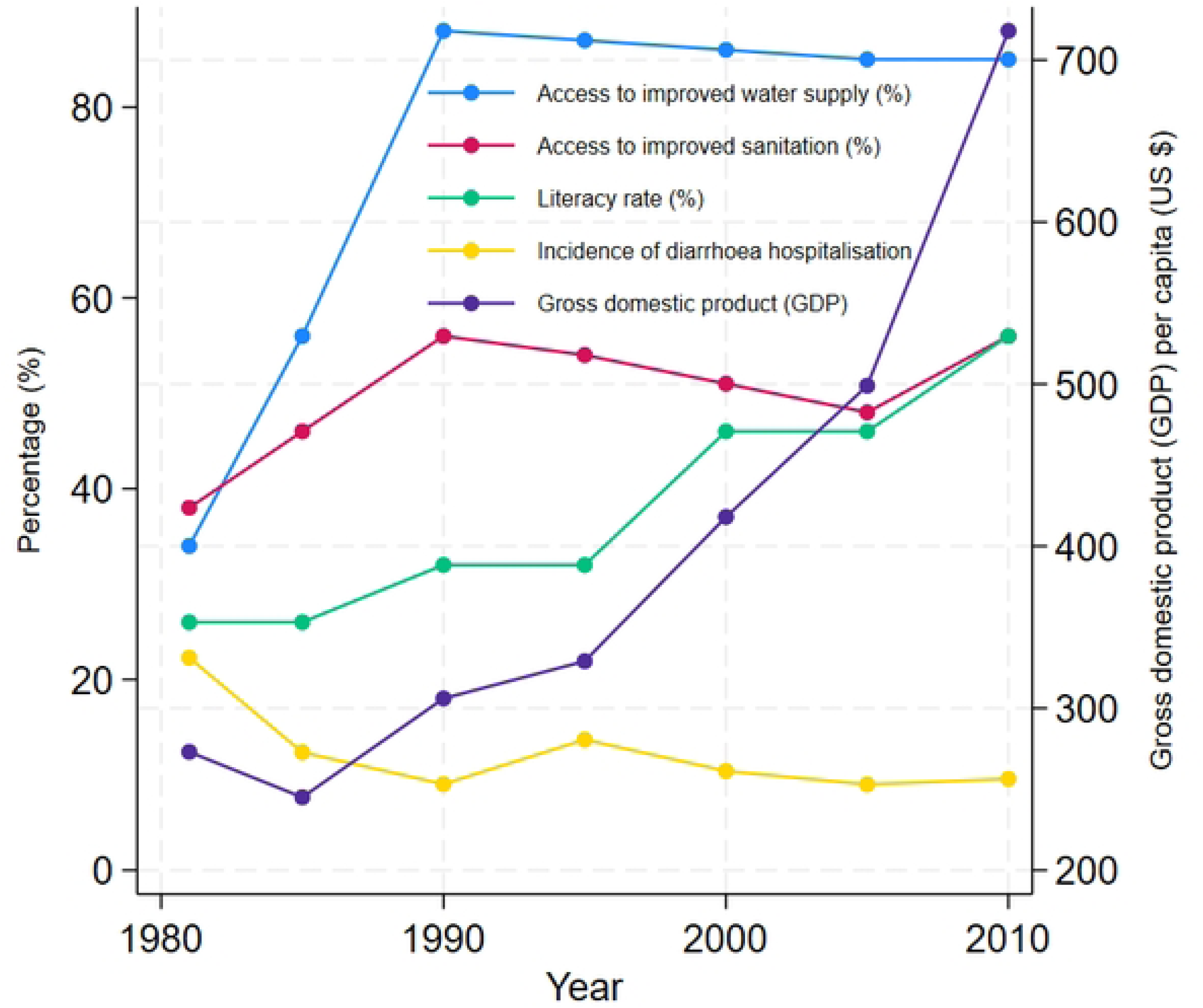
Demographic, economic, improved water supply access, sanitation coverage and diarrhoea hospitalisation trends in Dhaka, Bangladesh 1981-2010.

**Fig 4.**
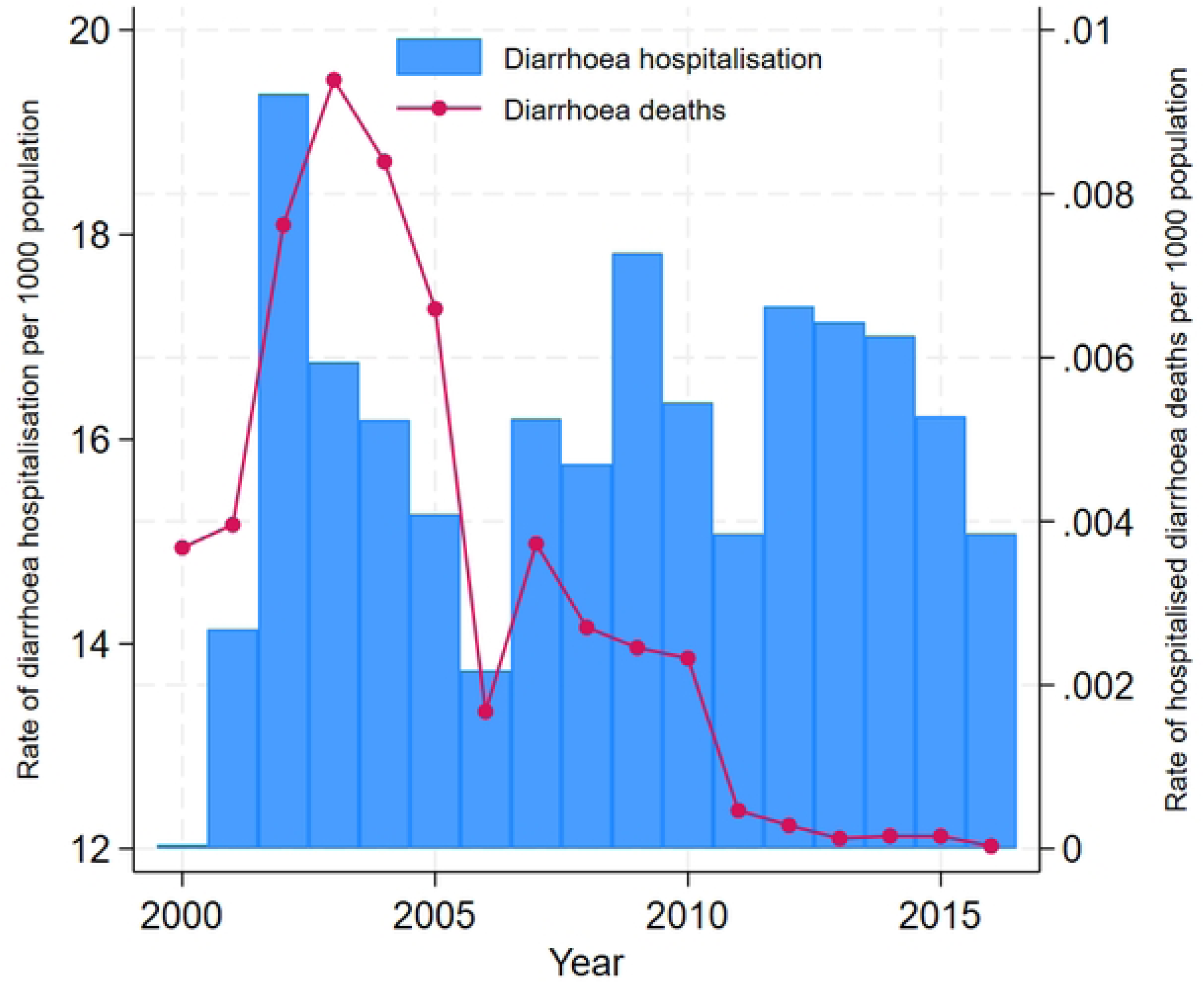
National trends of diarrhoea hospitalisation and deaths in Bangladesh during 2000-2016.

Figure 5 shows the seasonally adjusted relationship between temperature with relative risk of diarrhoea hospitalisation in all ages across multiple lags based on the DLLM model. There was a linearly increased risk of diarrhoea hospitalisation associated with temperature on the same day and a decreased risk at lag 1 day with negligible effects observed until lags of 28 days. The relationship with rainfall exhibited a broadly linear increased diarrhoea hospitalisation risk with heavy rainfall and the effect persisting for up to 8 days following exposure. We observed no increased risk of diarrhoea hospitalisation with low rainfall. In addition, relative humidity was inversely associated with diarrhoea hospitalisations on the same day with effects lasting up to 3 days. Using the DLLM, diarrhoea hospitalisations in all ages and ≤5 children in Dhaka were estimated to increase by 3.4% (95% CI: 3.0 – 3.7) and 3.9% (3.8 – 4.7) per 1°C increase in daily mean temperature, respectively (Table 1). The results did not change significantly during the sensitivity analyses (S2 File).

**Figure 5.**
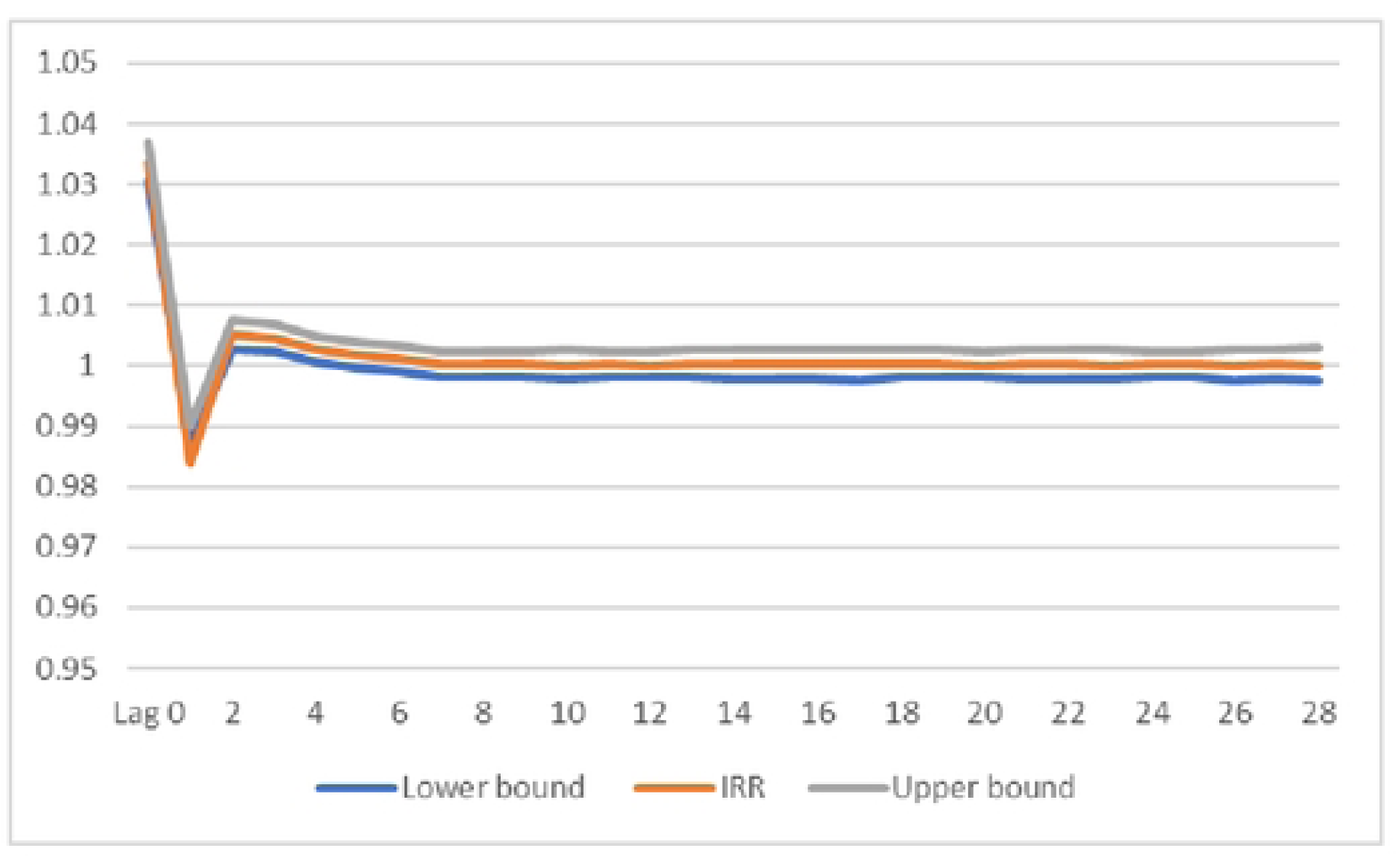
Lagged effects of ambient temperature on diarrhoea hospitalisations in all ages in Dhaka, Bangladesh.

**Table 1.**
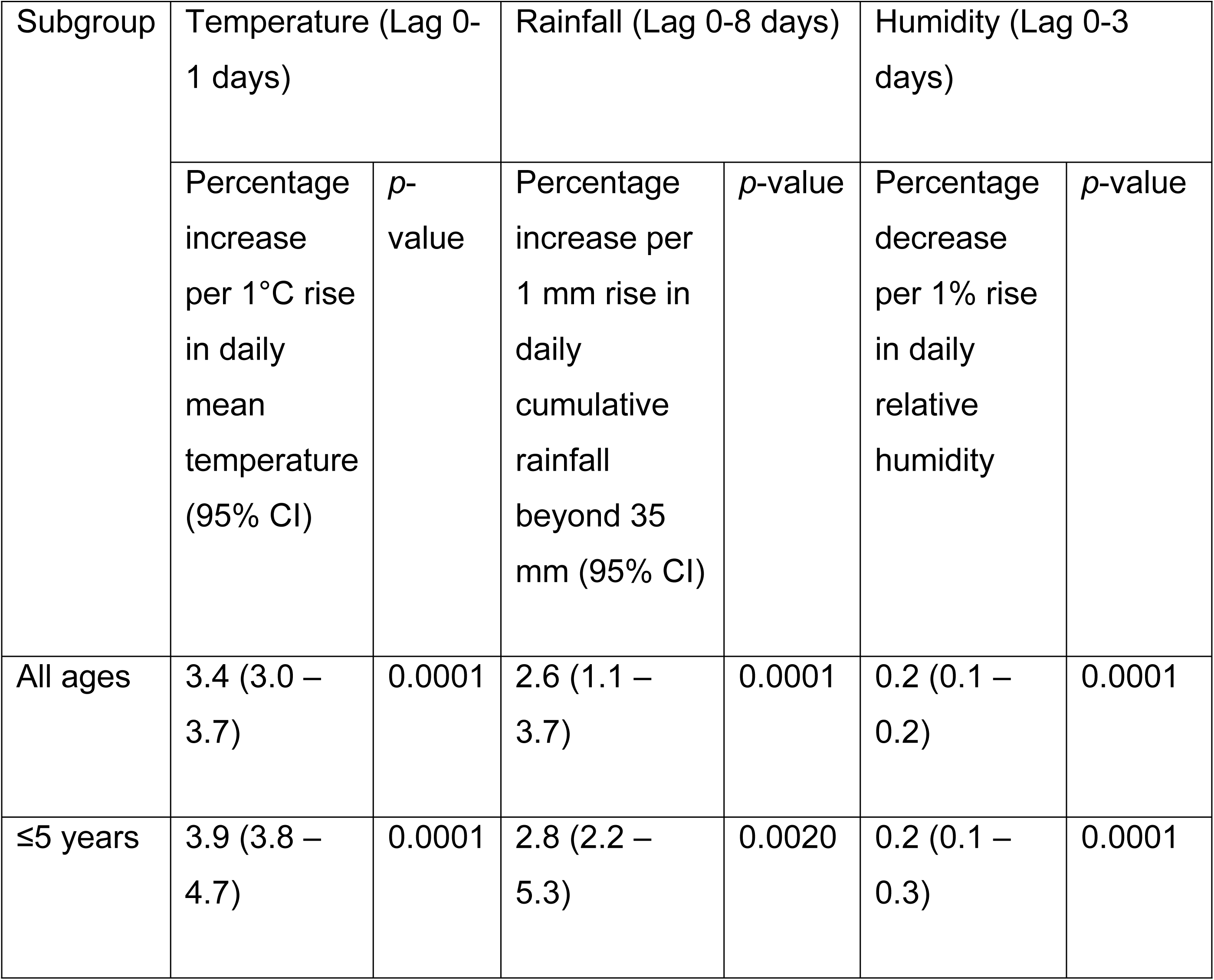
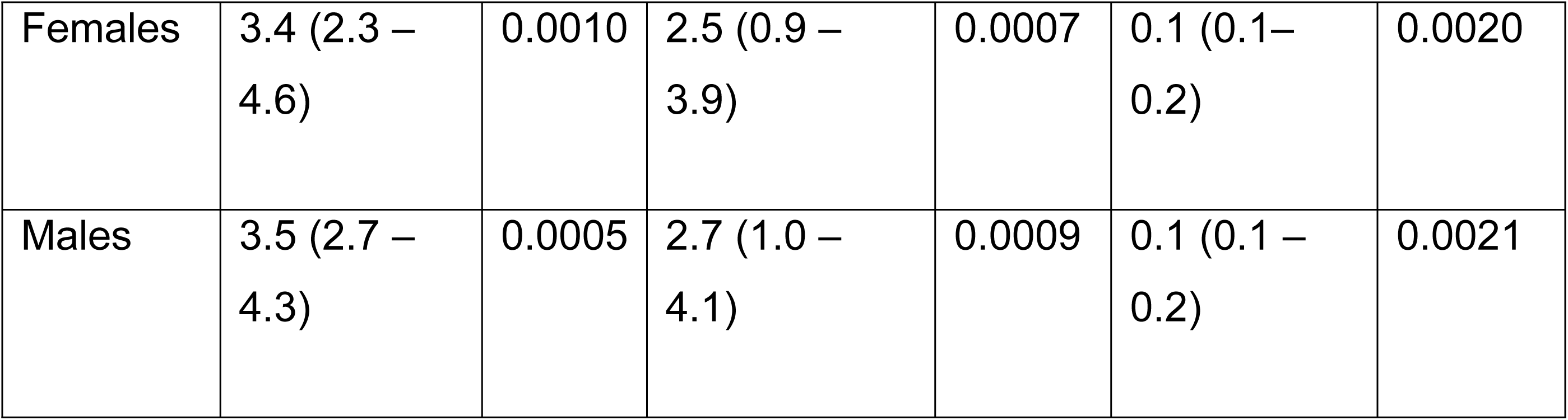
Percentage increase in diarrhoea hospitalisation associated with one unit increase in temperature, rainfall or humidity in Dhaka, Bangladesh.

| Subgroup | Temperature (Lag 0-1 days) |  | Rainfall (Lag 0-8 days) |  | Humidity (Lag 0-3 days) |  |
| --- | --- | --- | --- | --- | --- | --- |
|  | Percentage increase per 1°C rise in daily mean temperature (95% CI) | *p*-value | Percentage increase per 1 mm rise in daily cumulative rainfall beyond 35 mm (95% CI) | *p*-value | Percentage decrease per 1% rise in daily relative humidity | *p*-value |
| All ages | 3.4 (3.0 – 3.7) | 0.0001 | 2.6 (1.1 – 3.7) | 0.0001 | 0.2 (0.1 – 0.2) | 0.0001 |
| ≤5 years | 3.9 (3.8 – 4.7) | 0.0001 | 2.8 (2.2 – 5.3) | 0.0020 | 0.2 (0.1 – 0.3) | 0.0001 |
| Females | 3.4 (2.3 – 4.6) | 0.0010 | 2.5 (0.9 – 3.9) | 0.0007 | 0.1 (0.1– 0.2) | 0.0020 |
| Males | 3.5 (2.7 – 4.3) | 0.0005 | 2.7 (1.0 – 4.1) | 0.0009 | 0.1 (0.1 – 0.2) | 0.0021 |

As shown in Table 2, diarrhoea hospitalisation in all ages is predicted to increase by 5.1% and 7.4% under the 1.5°C and 2°C GWLs, respectively. Diarrhoea hospitalisation in children under 5 years of age may be increased by 5.9% (95% CI: 5.7 – 7.1) under the 1.5°C and by 7.8% (95% CI: 7.6 – 9.4) under the 2°C GWL. With a projected rise in Dhaka’s temperature by 4.3°C by the 2100, diarrhoea hospitalisation could rise by 15.9% and 20.2% in all ages and ≤5 children, respectively. Compared to all age groups, temperature increases under all scenarios showed higher impacts in children under 5 years of age. The impact of temperature increase on diarrhoea hospitalisation varied by gender with a slightly higher influence in males compared to females (5.1 – 6.8% Vs 5.8 – 7.0%) (Table 2). If the baseline relationship from 2010 between meteorological factors and diarrhoea hospitalisation risk and all other factors including population remain constant in the future, an increase in mean temperature by 0.91°C, 1.5°C, 2°C or 4.3°C could result in 4,760, 7,840, 10,360 and 22,260 additional diarrhoea hospitalisations annually in Dhaka Hospital by the 2100, respectively. Applying the same baseline risk as Dhaka, a temperature increase of 4.3°C by the 2100 could result in 385,955 additional hospitalisations across Bangladesh annually.

**Table 2.** Impacts of projected temperature increase on diarrhoea hospitalisation risk in Dhaka Bangladesh under various climate change scenarios.

| Subgroup | Relative risk (%) of diarrhoea hospitalisation |  |  |  |
| --- | --- | --- | --- | --- |
|  | With 1.5°C<br>GWL*<br>(95% CI) | With 2°C GWL<br>(95% CI) | With 0.91°C<br>predicted<br>increase by<br>2100 (95% CI) | With 4.3°C<br>predicted increase<br>by 2100 (95% CI) |
| All ages | 5.1 (4.5 – 5.6) | 6.8 (6.0 – 7.4) | 3.1 (2.7 – 3.4) | 14.6 (12.9 – 15.9) |
| ≤5<br>children | 5.9 (5.7 – 7.1 ) | 7.8 (7.6 – 9.4) | 3.6 (3.5 – 4.3) | 16.8 (16.3 – 20.2) |
| Females | 5.1 (3.5 – 6.9) | 6.8 (4.6 – 9.2) | 3.1 (2.1 – 4.2) | 14.6 (9.9 – 19.8) |
| Males | 5.3 (4.1 – 6.5) | 7.0 (5.4 – 8.6) | 3.2 (2.5 – 3.9) | 15.1 (11.6 – 18.5) |

\*GWL: Global Warming Level targeted in the Paris Agreement

## Discussion

While numerous studies have explored the effects of climate variability on diarrhoea worldwide [31], only a small number of studies have attempted to estimate the impacts of future climate change on diarrhoea [9, 10, 13]. Using empirical data from icddr,b Dhaka Hospital, our findings suggest significant baseline burden of acute diarrhoeal disease in Dhaka associated with ambient temperature after adjusting for the confounding effects of humidity, heavy rainfall, seasonal patterns, long-term trend, and day-of-the-week. This risk may aggravate further as climate continues to change. Our findings reveal that temperature-related diarrhoeal disease could increase by 7.4% in people of all ages and 9.4% in children under 5 years of age under the 2°C global warming level compared to the reference period of 1981 – 2010. This increase would be limited to 5.6% in people of all ages and 7.1% in children under 5 years of age if the 1.5°C global warming target could be reached. However, diarrhoeal disease hospitalisation could rise by up to 15.9% in all ages and 20.2% in children under 5 years if Dhaka’s temperature increases by 4.3°C by the 2100 as projected by various climate models [24].

With notable improvements in socio-economic condition and healthcare provision and advances in water, sanitation and hygiene infrastructure in Bangladesh, diarrhoea mortality in children has declined significantly from 15.1 per 1000 in 1980 to 6.0 per 1000 live births in 2015 [32, 33]. The rate of diarrhoea hospitalisation in Dhaka Hospital also suggests a declining trend. Despite success in reduction of diarrhoea mortality, outbreaks and hyperendemicity continue to plague the nation and diarrhoea remains a major cause of morbidity in Bangladesh for people in all age groups including children under 5 years of age [16, 34].

The baseline risk of diarrhoea hospitalisation with temperature in Dhaka observed in this study was smaller compared to many studies [31]. Given the high baseline rates, the temperature-attributable numbers of diarrhoea hospitalisation were still substantially large. In this study, the observed effects of temperature on diarrhoea hospitalisation were mostly immediate with negligible effects observed for up to 28 lag days. This could be linked to an acute mechanism of association, such as increased consumption of contaminated drinking water or increased food spoilage during hot weather [35].

In this study, diarrhoea hospitalisation decreased with mean temperature at the lag of 1-day following an initial increase in diarrhoea hospitalisation on the same day. In environmental epidemiological studies, short-term increase in risk of diarrhoea followed by a protective effect at a longer lag is known as ‘harvesting’ or ‘short-term displacement’ effect [29]. Prior studies eliciting the impact of climate parameters on diarrhoea rarely reported the occurrence of any harvesting effect. Aik et al (2019) examining climate variability on diarrhoea incidence in Singapore using time series analysis reported evidence of harvesting effect of diarrhoeal disease due to relative humidity six weeks later [36]. Another study investigating the influence of climate on diarrhoea in Vietnam reported evidence of harvesting effect due to rainfall but did not include the protective effect of rainfall into the final model [37].

Reductions in at-risk population following diarrhoea episodes likely contributed to the decreased diarrhoea hospitalisations observed the following day. Furthermore, family members and allies are likely to change their personal hygiene practices, food preparation behaviour and healthcare seeking practice following the hospitalisation of an affected member with an episode of diarrhoeal disease, which could also reduce their risk of suffering and/or hospitalisation for diarrhoea. While evidence of protective behavioural changes following a diarrhoea episode is scanty, a previous Norovirus outbreak report suggested that affected passengers of a cruise ship were three times less likely to indulge in risky behaviour including attending a casino, gym, on board events or shore side tours compared to unaffected passengers [38].

In this study, gender was not found to modify the temperature–diarrhoea relationship with males and females having nearly similar risk. However, the effect of mean temperature on diarrhoea was highest among children under 5 years of age. Compared to adults, young children may be more vulnerable to the effects of temperature owing to their immature immune system and low self-care capacity [39]. However, one global cross-sectional study did not find any significant relationship between temperature and diarrhoea in children under 5 years of age [40]. On the contrary, a study from Ethiopia reported significant effect of temperature on childhood diarrhoea [41]. Despite the potential vulnerability of children to climate change, empirical studies examining the role of age on the temperature–diarrhoea relationship are scarce. Future research examining the modulating impacts of age on climate–diarrhoea relationships can shed further light on this topic.

Our study has several strengths and limitations. The estimate of baseline impact of temperature on diarrhoea is based on high-quality, continuous data covering a 30–year period from the largest diarrhoea hospital in Dhaka, Bangladesh. In the context of a low- and middle-income country, the availability of such a dataset is rather exceptional. Most importantly, the analysis used temperature-diarrhoea relationship derived from a model that not only considered the effect of temperature on diarrhoea, but also the combined effects of temperature and other meteorological parameters including the immediate and lagged effects of humidity and rainfall. Furthermore, incorporation of WASH factors and other socio-economic indicators into the model during sensitivity analyses revealing comparable results further enhanced the confidence in our findings. Nevertheless, some limitations remain. Hospitalisations for diarrhoeal diseases at the icddr,b Dhaka Hospital may not be representative of all affected diarrhoea cases from Dhaka. It is believed that for every patient seeking hospital care for diarrhoea, there are four patients seeking care at the community [42, 43]. Hospital admissions, therefore, only represent a tip of the iceberg. Additionally, these represent the more severe cases. However, Dhaka Hospital is the largest diarrhoea treatment facility in the country and served a population of about one and a half million in 2010. Furthermore, icddr,b Dhaka Hospital services are likely to be accessed predominantly by the disadvantaged group and the surrounding slum dwellers. The estimates from this study are therefore conservative and likely to underestimate the actual effect of temperature variations on diarrhoeal disease transmission in Dhaka. Nonetheless, severe cases from the less privileged group are important given that such cases have implications for healthcare utilisation and may require policy changes to allocate more resources to improve diarrhoea hospital care in Dhaka. Although less severe cases and the elite class are less likely to be included, the results of this study comparing temperature-diarrhoea relationship over time are still valid.

Furthermore, climate change impact calculations were based on global warming targets of 1.5 - 2°C by the 2100. However, as evident from several models from Bangladesh, this rise in temperature is likely to be reached much sooner and the temperature of Dhaka could increase by 4.3°C by the 2100. Although climate models are prone to biases, we calculated the likely impacts based on a few local climate projections and found that diarrhoea hospitalisations in all ages could increase by up to 15.9% for Dhaka. In addition, we assumed that the baseline relationship between meteorological factors and diarrhoea hospitalisation risk will remain constant in the future. However, future diarrhoea risk will be affected by many other factors including population density, access to healthcare and vaccines that we did not aim to model since we wanted to quantify climate change risks only. Although the role of access to safe water and sanitation were accounted for in the temperature-diarrhoea relationship estimates during the sensitivity analyses, the possible effects of other adaptive health policies on diarrhoea including increased access to vaccines targeting cholera and rotavirus on diarrhoea prevalence and hospitalisation could not be incorporated in these models due to lack of available data. However, seasonal adjustment accounted for the variations to some extent. In the light of the above strengths and limitations, the predicted increase of 3.1 – 15.9% in diarrhoea hospitalisations with 0.91 – 4.3°C increase in temperature due to climate change should be considered indicative.

## Conclusions

This study demonstrated a statistically significant relationship between ambient temperature, relative humidity and heavy rainfall in the sub-tropical city of Dhaka. Given the high prevalence of diarrhoeal diseases in Bangladesh, even a small increase in risk stemming from increased environmental temperature in the context of climate change could lead to substantial burden and overwhelm the healthcare services in Dhaka. This underscores the importance of preparing the city for prevention and management of diarrhoeal diseases in anticipation of such increases in the future.

## Data Availability

According to institutional data policy of the icddr,b (International Centre for Diarrhoeal Disease Research, Bangladesh), only summary of data can be publicly displayed or can be made publicly accessible. To protect intellectual property rights of primary data, icddr,b cannot make primary data publicly available. However, upon request, Institutional Data Access Committee of icddr,b can provide access to primary data to any individual, upon reviewing the nature and potential use of the data. Requests for data can be forwarded to: Ms. Armana Ahmed, Head, Research Administration, icddr,b, Dhaka, Bangladesh, Phone: +88 02 9827001-10 (ext. 3200).

## Acknowledgments

This study was completed as a part of a PhD thesis by FH. The authors would like to thank the Commonwealth Scholarship Commission for funding the PhD study. The authors would also like to thank members of the Research Administration team of icddr,b, National Health Foundation and Research Institute (NHF&RI) and Bangladesh Meteorological Department (BMD) for their support to data collection.

## Supporting information

**S1 File. Additional information on health data.**

**S2 File. Sensitivity analyses.**

